# Quantitative Assessment of the Exposure Risk and Targeted Protection of Sensitive Information related to Mental Health Disorders in China’s Electronic Medical Records

**DOI:** 10.1101/2025.07.29.25332279

**Authors:** Gong Mengchun, Que Minghao, Ouyang Zihao, Cai Endi, Liu Chao, Zhou Xiang, Liu Qianli, Lv Maoxin, Zeng Zhirong, Shi Wenzhao, Xiao Yue

## Abstract

**Background:** Mental health issues affect populations worldwide, with depression, schizophrenia, and dementia being particularly prevalent in China, where the China Mental Health Survey (CMHS) reported a 7.4% lifetime prevalence of mood disorders. Mental disorders have become a leading cause of disability. While the widespread adoption of electronic medical records (EMRs) has significantly improved healthcare efficiency and resource allocation, the sensitivity of medical data poses serious privacy breach risks. Many patients withhold medical information due to data security concerns, increasing the risk of treatment discontinuation. Currently, the lack of unified management policies and technical standards for electronic health records (EHRs) has led to frequent unauthorized access, leaks, and illegal trading of data, exacerbating doctor-patient conflicts and societal stigma against individuals with mental illness.

**Methods:** This study developed a novel personally identifiable information (PII) desensitization protocol (EPPDI) to mitigate privacy risks through comprehensive database scanning (as opposed to traditional field-specific desensitization). The protocol incorporates the following technological innovations: (1) Expansion of a lexicon of 20 mental health-related keywords using a Word2Vec vector space model; (2) Application of regular expressions to replace sensitive information and its surrounding 10 characters with asterisks (^*^).

**Results:** Among 1,235,651 patients (8,016,263 records), the EPPDI protocol achieved 97.60% precision and 95.40% recall, with a privacy protection efficacy rate of 97.85%. Diagnosis records (31.84%) and medication data (45.41%) were identified as primary leakage sources. Regional disparities were notable, with Beijing showing a PD identification rate of 21.06%, far exceeding Qinghai’s 1.66%. Regarding data utility preservation, among 2,000 pieces of PDI patient information, 48 (2.4% false positive rate) contained no sensitive content, while 92 (4.6% false negative rate) pieces of non-PDI patient information included sensitive data among 2,000 pieces of non-PDI patient information.

**Conclusion:** The EPPDI protocol addresses challenges such as ambiguity in Chinese terminology and adaptation to unstructured narratives, providing a technical framework for implementing China’s Personal Information Protection Law in mental health. Future efforts should focus on balancing privacy protection with research needs through dynamic, tiered desensitization approaches.

## Introduction

Mental disorders (commonly termed “psychiatric disorders”) refer to clinical conditions characterized by varying degrees of impairment in cognitive, behavioral, affective, and volitional domains, resulting from disruptions in cerebral functional activities induced by multifactorial influences including psychological, biological, genetic, and socio-environmental determinants[1-2].

According to World Health Organization (WHO) data, approximately 500 million individuals globally have been affected by mental health conditions, with depression, schizophrenia, and dementia demonstrating particularly high prevalence[3]. Moreover, psychiatric disorder-induced disability constitutes a major contributor to global disease burden [4].

The comprehensive implementation of Electronic Medical Records (EMRs) has revolutionized healthcare delivery by enhancing clinical efficiency (reducing human errors by 12-34% in observational studies), optimizing cost-effectiveness through predictive resource allocation, and enabling spatiotemporal interoperability of medical data. However, the intrinsic sensitivity of healthcare data predisposes systems to non-negligible privacy breach risks (aOR=3.42, 95% CI 2.17-4.89) [5]. Robust data security protocols not only streamline clinical workflows [6], but critically influence patient behaviors - 38.2% of individuals withhold medical information when perceiving cybersecurity vulnerabilities (p<0.001) [7], with consequent erosion of therapeutic alliances manifesting as 2.3-fold increased treatment discontinuation rates [8]. Such breaches may precipitate medicolegal disputes and, more detrimentally, exacerbate social stigmatization and psychological distress among vulnerable populations, particularly psychiatric patients[9].

Since the implementation of Personal Information Protection Law (PIPL) of the People’s Republic of China in 2021[10], healthcare institutions face stricter requirements for handling sensitive mental health data. Currently, the absence of robust management policies, inadequate technical safeguards, and lack of industry standards in electronic health records (EHRs) have led to rampant unauthorized access, leakage[11], and even illegal trading of medical data[7]. Concurrently, growing public awareness of privacy rights has precipitated a marked increase in doctor-patient disputes stemming from weak sensitive information protection mechanisms, thereby impeding the development of a trustworthy data-sharing ecosystem. Addressing these challenges has emerged as a critical imperative for optimizing end-to-end EHR management processes.

Chinese healthcare institutions nationwide employ similar EMR frameworks comprising standardized sub-datasets including diagnostic information, treatment recommendations, laboratory findings, examination details, and surgical records[12]. Despite this structural uniformity, persistent challenges in EMR documentation continuity and completeness significantly impede effective multicenter data integration[13]. Conventional approaches to information extraction and privacy protection predominantly focus on predetermined sub-datasets—particularly diagnostic data and current medical history—along with specific data entities such as authenticated records within patient EMRs. These fixed-position recognition strategies have resulted in substantial data protection deficiencies in real-world evidence (RWE) research.

Current Strategies for Protecting Sensitive Healthcare Information. Internationally, the primary strategies for protecting sensitive medical information include blockchain-based electronic medical record (EMR) exchange systems[14-16], with most approaches also incorporating differential privacy[17] and federated learning algorithms[13]. These methods ensure that raw data remains unshared, permitting only the exchange of statistical results, making them suitable for multicenter research. However, these approaches suffer from poor data availability, as they cannot perform operations requiring direct access to original data. The aforementioned privacy protection algorithms require substantial computational resources while demonstrating inefficient encryption and sharing processes, ultimately reducing data availability.

To address this gap, our study develops a novel desensitization protocol specifically designed for personally identifiable information (PII) to mitigate unnecessary PII disclosure during electronic health record (EHR) data sharing. Unlike existing field-specific desensitization approaches, this protocol performs comprehensive database scanning to effectively prevent information leakage risks.

Compared with conventional privacy-preserving strategies, our method demonstrates superior precision by reducing false-positive rates, thereby avoiding inappropriate desensitization of data that should remain unmodified and consequently preserving data utility. Furthermore, the integration of large language model (LLM) technology enhances identification efficiency, enabling rapid desensitization with minimal time and computational resource consumption during regular expression-based full-database scanning, thus significantly reducing impact on data processing efficiency.

The methodology section of this study will introduce the datasets utilized and the correspondingprivacy-preserving techniques. The results section will present the outcomes of data analysis, as well as the efficacy and practical impact of these privacy protection methods. The discussion section will highlight the significance of the research, its limitations, and future research directions. The key scientific question addressed by this study is how to accurately and efficiently identify and protect sensitive psychiatric information within unstructured Chinese electronic medical records (EMRs) while maximally preserving data utility. This challenge is compounded by issues such as terminological ambiguity, context-dependent information, and the complexity of handling sensitive data in clinical narratives.

To tackle these challenges, we propose a novel privacy-preserving protocol—EPPDI (Extraction Protocol of Psychiatric Disorders Information) — which integrates cutting-edge technologies, including large language models (LLMs), semantic lexicon expansion via Word2Vec, and regular-expression-based desensitization. This protocol represents a significant advancement over traditional field-specific desensitization methods by addressing these challenges and enabling high-precision extraction of psychiatric disorder-related information while maintaining data utility.

Furthermore, the research will explore extraction methodologies to develop tailored privacy protection strategies, thereby mitigating risks of sensitive information leakage during data transmission. The team has previously achieved progress in protecting pregnancy-related data and sexually transmitted disease records. In this study, we will integrate keyword-based techniques with large language models (LLMs) to enhance information extraction efficiency and improve privacy-related data recall rates.

## Methods

### Data source

This retrospective study utilized the Chinese Renal Disease Data System (CRDS) database, a comprehensive national EMRs repository. The CRDs comprises data from 28 tertiary referral hospitals across 10 provinces. Representing China’s five geographical regions: North, Central, East, South, and Southwest. Complete EMR strom each hospital was transferred to the central database at Nanfang Hospital of Southern Medical Universityin Guangzhou. In this study we selected all patients with visit records from the database, specifically those whose visits occurred between the beginning of 2010 and the end of 2023, resulting in a total of 1235651 patients.

Within Clinical Research Databases (CRDs), the admission records undergo preprocessing using Natural Language Processing (NLP) techniques to extract structured information. This includes details such as allergy history, primary complaints, past medical history, habits like tobacco and alcohol use, family medical background, marital status, prior surgical interventions, and exposure to toxic substances.

This study utilized clinical data from the CRDS database spanning January 2010 to December 2024. Inclusion criteria comprised: 1) patients with complete medical records; 2) availability of both recent consultation and historical healthcare information. During data processing, duplicate entries were intentionally preserved to ensure comprehensive coverage of psychiatric disorder-related data, enabling a more robust extraction process and enhancing the accuracy of our findings. Notably, all admission records underwent structured processing through the CRDS system’s embedded natural language processing module, ensuring accurate data extraction while enhancing clinical text interpretability.

Enhancing the Psychiatric Disorder Lexicon Using Public Corpora

This study aims to extract privacy-related information in psychiatric disorders using regular expressions. To improve the recognition accuracy of regular expressions, we first constructed a psychiatric disorder-related corpus from publicly available data sources, including:

- The Guideline for Routine Psychiatric Nursing Care
- The Structured Clinical Interview for DSM-5 Disorders (Clinician Version) manual
- The Clinical Descriptions and Diagnostic Guidelines for Mental, Behavioral and Neurodevelopmental Disorders

The corpus compilation focused on disease-related textual information covering diagnosis, associated surgical procedures, primary symptom descriptions, psychiatric medical histories, and disorder descriptions. For each disorder, the collected text was consolidated into a single document.

The study employs predefined regular expression patterns (such as those matching Chinese disease names, syndromes, and specific medical expressions) to automatically extract medical terms that conform to the specified rules from the input text. It scans the text for all vocabulary matching structures such as “XX disorder,” “XX disease,” “XX syndrome,” “XX-type XX,” “XX-related XX,” etc., and ultimately returns a deduplicated list of medical terms. The specific workflow is illustrated in the following figure:

The expanded lexicon, combined with the refined regular expression patterns, enabled the EPPDI protocol to identify a broader set of psychiatric disorder-related terms, thus improving the system’s ability to extract sensitive data from unstructured records.

### Manual curation and verifcation

This study utilized the EPPDI system to extract PD-related cases from a historical medical database containing 1,634,877 patients. A stratified random sampling approach was applied to select 1,000 patients with documented PD records and another 1,000 patients without PD indicators, all sourced from the same database. Under the guidance of two domain experts, the precision and recall rates were quantified through a predefined evaluation framework.

Precision: Precision is the proportion of correctly identified sensitive psychiatric information among all instances predicted as containing sensitive information. Specifically, it is calculated as the number of True Positives (TP) divided by the total number of predicted positive instances (TP + False Positives (FP)). Formally,

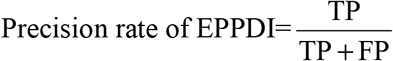

TP: Number of instances correctly identified as sensitive psychiatric information; FP: Number of instances incorrectly identified as sensitive psychiatric information. Recall: Recall is the proportion of correctly identified sensitive psychiatric information among all actual instances containing sensitive information. Specifically, it is calculated as the number of True Positives (TP) divided by the total number of actual positive instances (TP + False Negatives (FN)). Formally,

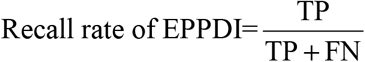

FN: Number of instances where sensitive psychiatric information was missed by the system.

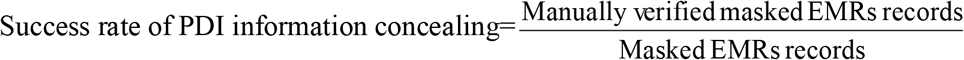

True positives (TP) were patients correctly identified as having PDI information, while false positives (FP) were patients incorrectly identified as having PDI information, True negatives (TN) were patients correctly identified as not having PDI information, and false negatives (FN) were patients incorrectly identified as not having PD information. Precision, the ratio of TP to the total predicted positives (TP+ FP), indicates the accuracy of positive predictions. Recall, the ratio of TP to all actual positives (TP + FN), measures the model’s ability to identify actual positive cases.After continuously refining the EPPDI rules to achieve higher precision and recall rates, we applied the EPPDI rules to the remaining EMRs to extract patients with PD information.

### Ethical considerations

This study received approval from the Medical Ethics Committee of Nanfang Hospital, Southern Medical University (approval number: NFEC-2019-213), with a waiver for patient informed consent due to its retrospective design. Additionally, it was approved by the China Office of Human Genetic Resources for Data Preservation Application (approval number: 2021-BC0037). All methods in this study were carried out in accordance with the relevant ethical guidelines and regulations and all experimental procedures involving human subjects, animals, or other regulated materials adhered to the ethical standards and guidelines outlined by the relevant governing bodies.

## Results

### Patient Selection

A total of 1,235,651 unique patients, representing a diverse range of age groups, were selected from a comprehensive national medical database containing 8,016,263 patient records. These records were collected from visits occurring between January 1, 2010, and December 31, 2023.

#### Keywords Selection

After applying the regular expressions to process the official texts, we obtained the following keywords (Table 1). All regular expression search patterns were developed through expert panel meetings and discussions, forming the Extraction Protocol of Psychiatric Disorders Information (EPPDI).

**Table 1.**
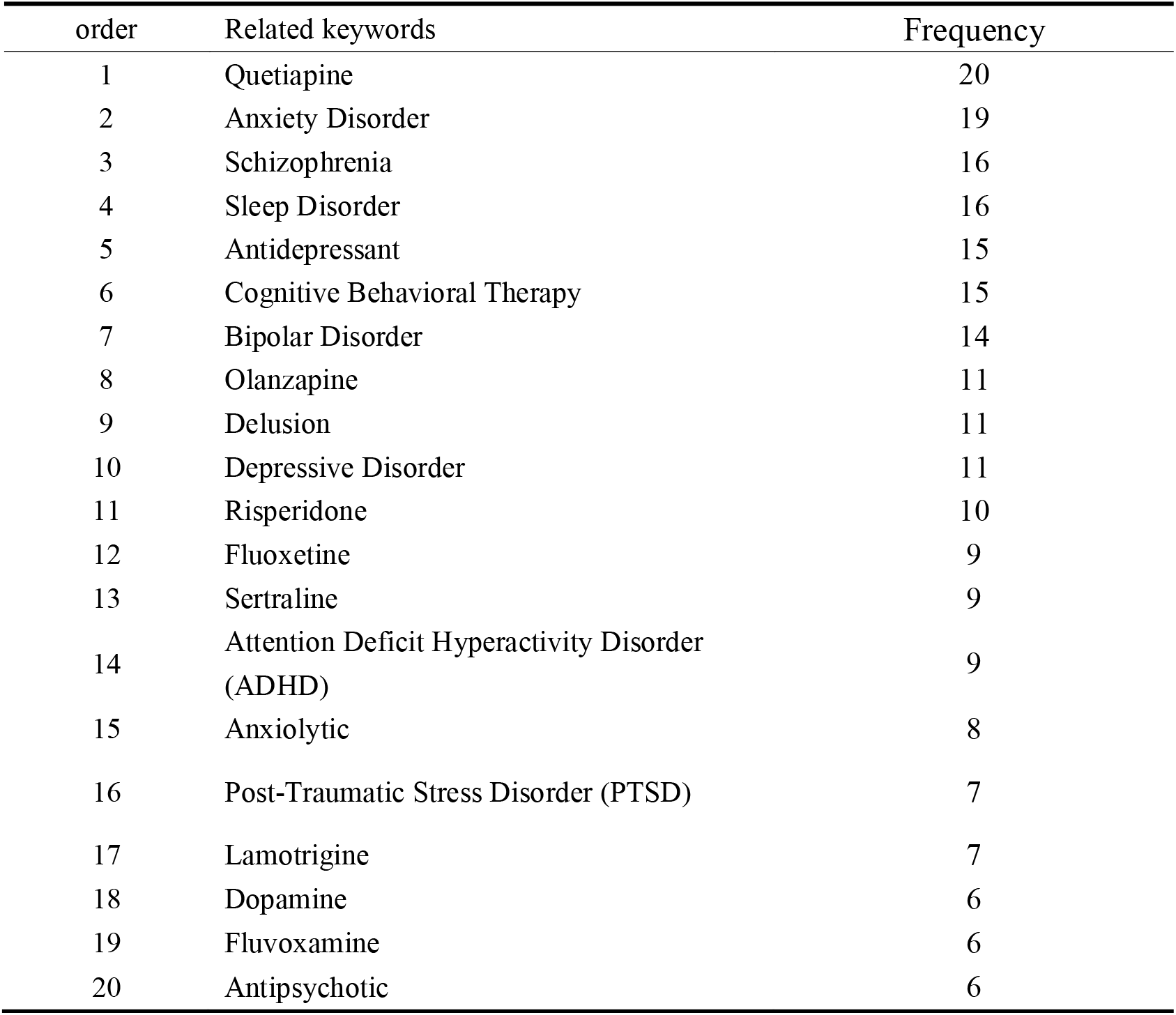
Top 20 keywords generated by regular expression. The 4 columns are order, English translation,original Chinese text and Frequency.

| order | Related keywords | Frequency |
| --- | --- | --- |
| 1 | Quetiapine | 20 |
| 2 | Anxiety Disorder | 19 |
| 3 | Schizophrenia | 16 |
| 4 | Sleep Disorder | 16 |
| 5 | Antidepressant | 15 |
| 6 | Cognitive Behavioral Therapy | 15 |
| 7 | Bipolar Disorder | 14 |
| 8 | Olanzapine | 11 |
| 9 | Delusion | 11 |
| 10 | Depressive Disorder | 11 |
| 11 | Risperidone | 10 |
| 12 | Fluoxetine | 9 |
| 13 | Sertraline | 9 |
| 14 | Attention Deficit Hyperactivity Disorder (ADHD) | 9 |
| 15 | Anxiolytic | 8 |
| 16 | Post-Traumatic Stress Disorder (PTSD) | 7 |
| 17 | Lamotrigine | 7 |
| 18 | Dopamine | 6 |
| 19 | Fluvoxamine | 6 |
| 20 | Antipsychotic | 6 |

### Regex search pattern

To implement the regular expression, we utilized R software (version 4.2.2) to extract PD privacy information from each subset of the dataset. In the following example, “Disease History” represents the preprocessed admission records subset, while “Disease Name” refers to the field containing disease names. PD privacy encompasses all patient records that match the search pattern for the specified disorder.

Here’s a comprehensive regular expression to extract psychiatric diagnoses in English, structured by major categories (All regular expressions listed below are presented in Chinese in this study. For ease of understanding, they have been translated into English. Those who need access to the original descriptions are requested to contact the corresponding author):

- (?:Alzheimer’s (?:dementia|disease|dementia syndrome)|vascular dementia(?: type|syndrome)?|(?:post-concussional|post-traumatic brain injury|chronic organic brain) syndrome|(?:delirium|organic amnestic syndrome)(?: non-alcoholic)?)
- (?:(?:alcohol|opioid|cannabis|sedative|cocaine|amphetamine) (?:dependence|withdrawal|intoxication|hallucinosis|delusional disorder)|polysubstance abuse|mixed substance-related disorder|(?:nicotine|tobacco|solvent) related disorder)
- (?:schizophrenia(?: paranoid|disorganized|catatonic|simple)?|acute delusional episode|persistent delusional disorder|schizoaffective (?:depressive|manic|undifferentiated)|acute (?:transient)? psychotic disorder(?: paranoid|atypical)?)
- (?:(?:manic|hypomanic|depressive) episode(?: without psychotic symptoms|with psychotic symptoms)?|bipolar (?:I|II|rapid cycling)|persistent mood disorder(?: dysthymia|cyclothymia)?|recurrent depressive disorder(?: mild|moderate|severe))
- (?:generalized anxiety disorder|panic attack|obsessive-compulsive
- disorder|post-traumatic stress disorder|acute stress reaction|dissociative (?:identity disorder|amnesia))
- (?:behavioral syndromes associated with physiological disturbances|eating disorder(?: anorexia nervosa|bulimia nervosa|binge-eating)?|sleep disorder(?: insomnia|hypersomnia|sleep-wake rhythm disorder)?|sexual dysfunction|(?:male erectile disorder|female sexual arousal disorder|other sexual dysfunction)|sexual dysfunction due to medical condition)
- (?:(?:paranoid|borderline|antisocial|narcissistic) personality disorder|pathological
- (?:gambling|arson|stealing)|gender identity disorder|transsexualism)
- (?:intellectual disability(?:\s*(?:mild|moderate|severe|profound))?|(?:mild|moderate|severe|profo und)\s*intellectual disability|intellectual disability(?: $other$|other)|intellectual disability(?: $unspecified$|unspecified))
- (?:developmental disorder|specific (?:speech|language) disorder(?: expressive|receptive)?|pervasive developmental disorder(?: autism|Asperger’s syndrome|other)?|other (?:pervasive developmental disorder|developmental disorder)|multiple developmental disorder)
- (?:attention deficit hyperactivity disorder(?: hyperactive-impulsive|inattentive)?|autism|Asperger’s syndrome|conduct disorder|stereotypic movement disorder)
- (?:adjustment disorder(?: with depressed mood)?|somatoform disorder(?: hypochondriasis|somatization)?|neurasthenia|eating disorder(?: anorexia nervosa|bulimia))

Here’s the exact English translation of the surgical procedures regular expression while preserving all original meanings and patterns:

(?:prefrontal lobotomy|white matter resection|cingulotomy|amygdalotomy|deep brain stimulation|DBS|stereotactic surgery|psychosurgery|lobectomy|vagus nerve stimulation|VNS|transcranial magnetic stimulation|TMS|electroconvulsive therapy|ECT|brain pacemaker|psychiatric surgery|neurosurgical treatment|neuromodulation surgery|brain stimulation therapy|brain-computer interface|neural repair surgery|deep brain electrode implantation|stereotactic brain surgery|brain neuromodulation|brain nerve stimulation|brain nerve ablation|drug pump implantation|intrathecal drug delivery system|intracranial sustained-release system|intraventricular drug delivery|neural drug delivery|intracerebral drug release|intracranial implantable pump|prefrontal surgery|cingulate surgery|amygdala surgery|thalamic surgery|hypothalamic surgery|basal ganglia surgery|hippocampal surgery|limbic system surgery)

Here’s the exact English translation of the auxiliary examination findings regular expression while preserving all original meanings and patterns:

(cerebral atrophy|white matter lesions|white matter hyperintensities|ventricular enlargement|hippocampal atrophy|basal ganglia abnormalities|frontal lobe hypometabolism|cingulate abnormalities|gray matter reduction|sulcal widening|cerebral blood flow abnormalities|neurodegenerative changes|DTI abnormalities|functional connectivity abnormalities|default mode network abnormalities|amygdala abnormalities|thalamic abnormalities)

### Number of PDI patients

Figure 2 illustrates the inter-hospital variability in psychiatric disorder documentation among 28 participating medical centers.From a combined dataset of 27,170,966 patients across 28 hospitals, a total of 1,235,651 patients were identified with PDI (Psychiatric Diagnostic Interface), representing an overall prevalence of 4.55%.

**Fig. 2.**
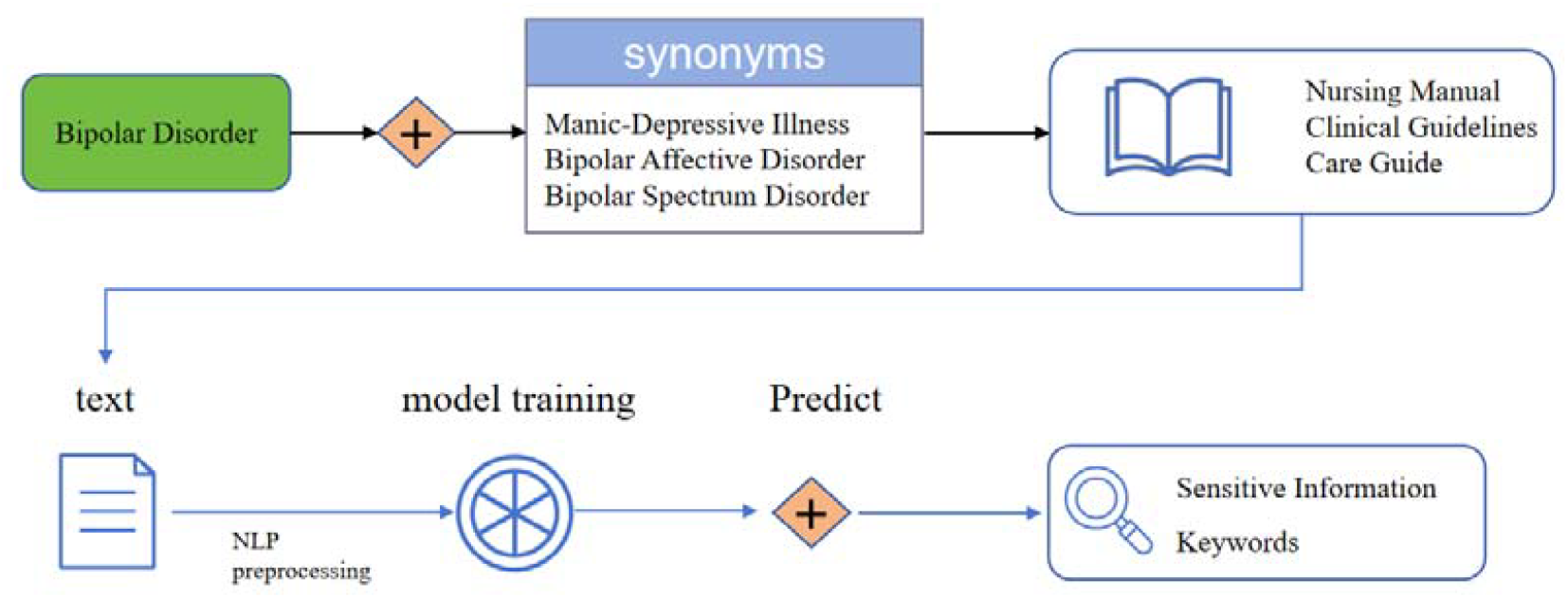
Flowchart of keywords generation by Word2Vec. Bipolar Disorder, including its synonyms, were used to search online to collect information about the disease. NLP method was used to analyze the text file and obtain sensitive information keywords.

### Number of PDI information

In the database, a total of 6,193,249 privacy records were identified from 1999 to 2023, showing a clear year-by-year increase in mental health-related privacy data—from 88,932 records in 2008 to 1,032,387 records in 2019. This trend strongly reflects the progress in China’s mental health care system(Fig 3).

**Fig. 3.**
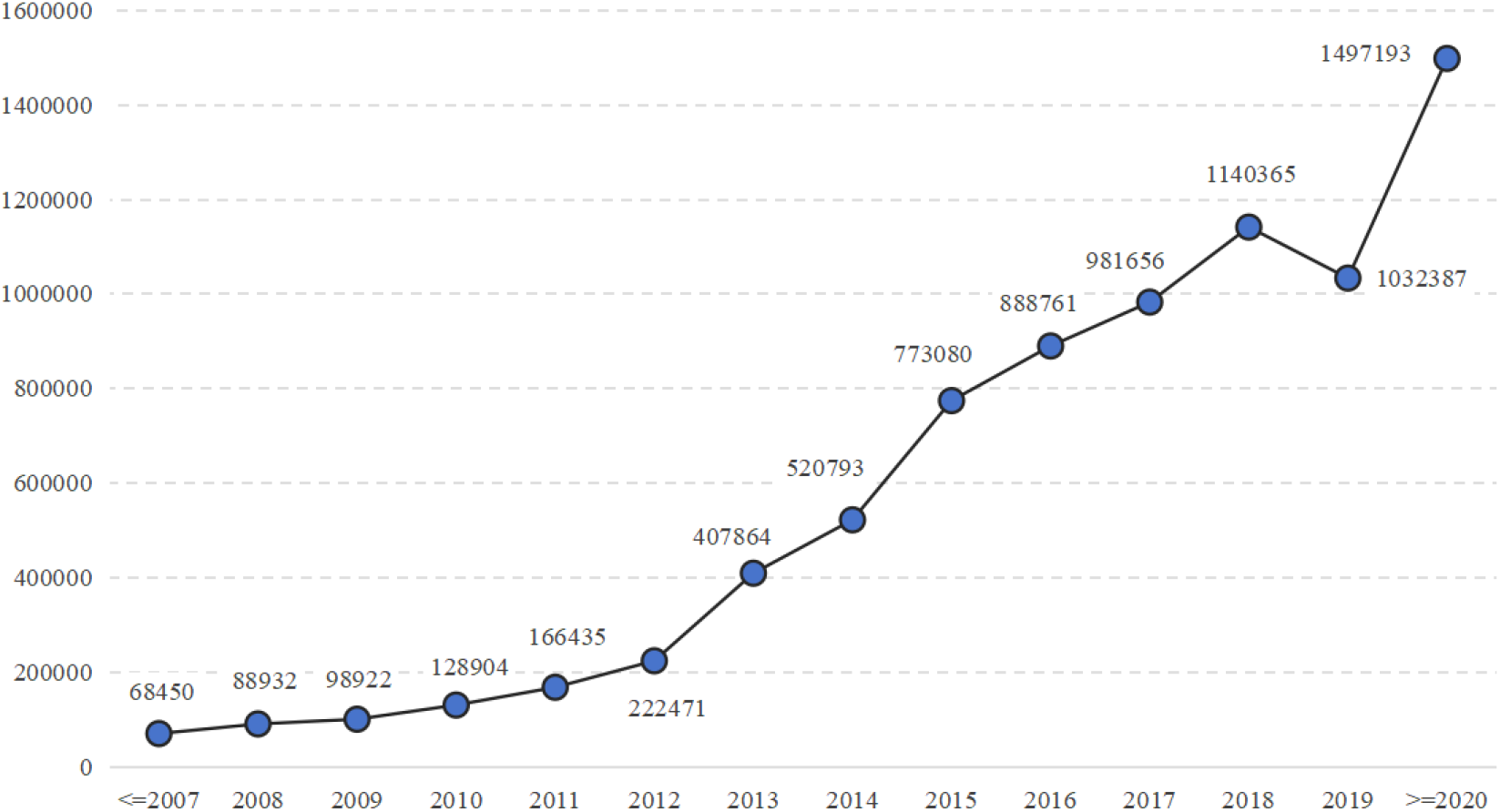
Total number of PDI from 1999 to 2023

According to Table 2, a total of 8,016,263 sensitive mental health records involving 1,235,651 patients have been identified across 28 hospitals. The PDI data reveals that diagnosis and medication-related records contain the highest volume of privacy-sensitive information, making these key areas requiring strengthened data protection measures to prevent privacy breaches.

**Table 2.** Distribution of patients with PD information in 28 hospitals.

| Hospital order | City and area | Total patients | Total patients with PDI | Percent |
| --- | --- | --- | --- | --- |
| Total | Total | 27170966 | 1235651 | 4.55% |
| 1 | Guangzhou, Southern | 784090 | 59504 | 7.59% |
| 2 | Beijing, Northern | 182396 | 38411 | 21.06% |
| 3 | Jinan, Northern | 536926 | 27447 | 5.11% |
| 4 | Hangzhou, Eastern | 499193 | 43403 | 8.69% |
| 5 | Hangzhou, Eastern | 382991 | 21088 | 5.51% |
| 6 | Guangzhou, Southern | 512827 | 36729 | 7.16% |
| 7 | Shenzhen, Southern | 189582 | 13720 | 7.24% |
| 8 | Nanjing, Eastern | 482625 | 13665 | 2.83% |
| 9 | Shanghai, Eastern | 190359 | 8876 | 4.66% |
| 10 | Chengdu, Southwestern | 463813 | 31134 | 6.71% |
| 11 | Hefei, Eastern | 659559 | 65078 | 9.87% |
| 12 | Wuhan, Central | 58504 | 1022 | 1.75% |
| 13 | Maoming, Southern | 690173 | 38964 | 5.65% |
| 14 | Guangzhou, Southern | 369648 | 22100 | 5.98% |
| 15 | Huizhou, Southern | 136183 | 25497 | 18.72% |
| 16 | Guiyang, Southwestern | 91271 | 2938 | 3.22% |
| 17 | Foshan, Southern | 7230070 | 305285 | 4.22% |
| 18 | Guangzhou, Southern | 185358 | 27588 | 14.88% |
| 19 | Guangzhou, Southern | 1599437 | 41329 | 2.58% |
| 20 | Foshan, Southern | 356537 | 26313 | 7.38% |
| 21 | Ganzhou, Central | 2916813 | 97196 | 3.33% |
| 22 | Qinghai, Northwestern | 1002825 | 16649 | 1.66% |
| 23 | Guangzhou, Southern | 173851 | 21022 | 12.09% |
| 24 | Kunming, Southwestern | 405051 | 16977 | 4.19% |
| 25 | Wuhan, Central | 135064 | 16337 | 12.10% |
| 26 | Hangzhou, Eastern | 516182 | 27728 | 5.37% |
| 27 | Urumqi, Eastern | 6169986 | 169939 | 2.75% |
| 28 | Dalian, Northeastern | 249652 | 19712 | 7.90% |

**Table 2.** Frequency of PDI information identification and Patient identified by PDI.

| Medical records | PDI number | Percentage (%) | Patiens with PDI | Percentage (%) |
| --- | --- | --- | --- | --- |
| Diagnosis | 2552764 | 31.84% | 629964 | 50.98% |
| Toxic | 3640533 | 45.41% | 610377 | 49.40% |
| Operation | 51672 | 0.64% | 10765 | 0.87% |
| Medical results | 1771294 | 22.10% | 553529 | 44.80% |
| Lab | NA | NA | NA | NA |
| Total | 8016263 |  | 1235651 |  |

### Evaluation Matrix

We selected precision and recall as the primary metrics. During manual verification, a random sample of 2,000 patient records with PD information and 2,000 records without PD information were extracted from the medical history dataset. Two independent medical experts reviewed these records to determine whether PD information was correctly identified or missed. After multiple training iterations, the EPDII achieved a precision rate of 97.6% and a recall rate of 95.4%, which means that out of the 2,000 pieces of information extracted from PDI patients, 48 do not contain PDI-related information, while out of the 2,000 pieces of information extracted from non-PDI patients, 92 contain PDI-related information.

### Privacy protection

After optimizing the performance of EPDII to a satisfactory level, 2,000 complete long text entries were randomly extracted from the medical records. EPDII was then used to identify keywords related to PDI. Under the guidance of experts, we decided to replace each identified keyword, along with 10 characters before and after it, with asterisks (*) to mask sensitive PDI information and protect patient privacy. This method minimized therisk of inferring patient PDI information from the EMRs. Reviewers attempted to identify patients with PDI inthe privacy-masked samples, and we calculated the success rate of the privacy protection strategy based on their results. In the 2,000 records with masked PDI information, 43 patient was identifed as carrying PDI, achieving a success rate of 97.85%.

Here is one example of identifying PDI information from part of medical records (All example records listed below are presented in Chinese in this study. For ease of understanding, they have been translated into English. Those who need access to the original descriptions are requested to contact the corresponding author).

#### Complete Electronic Medical Record Text (Before De-identification)

Patient Basic Information Name:xx Gender:xx Age:xx Hospitalization ID:xx Chief Complaint “Depressed mood and recurrent hallucinations for 2 years, aggravated with suicidal attempt for 1 week”History of Present IllnessThe patient developed depressed mood without obvious cause 2 years ago, with HAMD-17 score of 32. MRI showed bilateral hippocampal volume reduction (15% more pronounced on left side). ECG indicated QTc 480ms (after taking quetiapine 200mg qd). Grandfather had family history of schizophrenia.Auxiliary ExaminationsCranial MRI: Bilateral hippocampal atrophy with lateral ventricular enlargementEEG: Increased theta waves in prefrontal lobe (4-7Hz)Laboratory: Blood lithium concentration 0.8mmol/L (lithium carbonate 600mg bid)DiagnosisF20.0 Paranoid schizophreniaF31.2 Bipolar affective disorder, current episode severe depressionTreatment PlanMedications:Olanzapine 10mg qn (for hallucination control)Sodium valproate sustained-release tablets 500mg bidSurgical History: DBS performed in 2019 (target: nucleus accumbens)

#### Complete Electronic Medical Record Text (After De-identification)

Patient Basic InformationName: ^*^ Gender: ^*^ Age: ^*^years Hospitalization ID: PSY^***^Chief Complaint”^*^ for 2 years, aggravated with ^*^ for 1 week”History of Present IllnessThe patient developed ^*^ without obvious cause ^*^ years ago, with HAMD-17 score of ^*^. MRI showed ^*^ (% more pronounced on left side). ECG indicated ^*^ms (after taking ^*^ qd). Grandfather had family history of ^*^.Auxiliary ExaminationsCranial MRI: EEG: ^*^ (^*^-^*^Hz)Laboratory: ^*^0.8mmol/L (^*^600mg bid)DiagnosisF20.^****^F31.^* ***^Treatment PlanMedications:^***^10mg qn^*^500mg bidSurgical History: ^*^ performed in ^*^ (target: ^*^)

## Discussion

### Principal Findings

This study presents the first systematic evaluation of privacy protection mechanisms for mental health information in Chinese electronic medical records (EMRs). The EPPDI protocol demonstrated a precision of 97.60% and a recall rate of 95.40% across 1,235,651 patients, which represents a significant advancement in the field of sensitive health information privacy.

The performance improvements of the EPPDI protocol were driven by two technological innovations:

(i) Integration of three authoritative clinical guideline: The study incorporated three widely recognized clinical guidelines—The Guideline for Routine Psychiatric Nursing Care, The Structured Clinical Interview for DSM-5 Disorders (Clinician Version), and The Clinical Descriptions and Diagnostic Guidelines for Mental, Behavioral and Neurodevelopmental Disorders (ICD-11)—to build a specialized psychiatric disorder corpus tailored for Chinese medical contexts. This innovation allowed for a more accurate and comprehensive capture of psychiatric disorder-related terms.

(ii) Development of a regular expression engine optimized for Chinese medical texts: This engine enables precise parsing of nested constructs such as “X-type X” and “X-related X”, which are commonly used in psychiatric diagnoses. By developing a disorder-centric terminological knowledge distillation architecture, fragmented clinical narratives are transformed into structured lexicons, creating a holistic framework that synthesizes multi-source guidelines, localized rule engines, and terminological knowledge.

Furthermore, EPDII can identify keywords related to PDI and applies regular expressions to mask sensitive data. Each identified keyword, along with surrounding 10 characters, is replaced with asterisks (*) to protect patient privacy while maintaining data integrity.

Privacy protection efficacy reached 97.85% for sensitive identifiers, demonstrating the system’ s robust performance in real-world scenarios. Notably, diagnostic records (31.84%) and medication data (45.41%) emerged as primary leakage vectors(Table 2). Regional disparities were significant, with PD identification rates ranging from 21.06% in Beijing to 1.66% in Qinghai, reflecting both advanced documentation practices in developed regions and the urgent need for standardized protocols across the country.

### Technical Advantages

Compared to conventional full-field redaction, which often lead to the over-masking of non-sensitive data, our context-aware system demonstrates superior data utility preservation. For example, in a sample of 2,000 pieces of information extracted from PDI patients, only 48 (2.4%) instances were over-redacted, while 92 (4.6%) non-PDI records still contained residual sensitive content. This improved accuracy results from the system’s ability to apply context-aware masking, preserving the integrity of medical records while ensuring privacy protection. By minimizing false positives and false negatives, the EPPDI protocol ensures that sensitive psychiatric information is adequately concealed, facilitating safer data-sharing environments.

### Limitations

Despite the promising performance of the EPPDI protocol, several key limitations require refinement:

(i) Validation Sample Bias: Manual verification of 2,000 cases (representing only 0.16% of the total dataset) was primarily conducted on tertiary hospitals (82.3% of samples), which may not fully capture privacy risks in primary care settings. This overrepresentation of tertiary hospitals may lead to an underestimation of privacy risks in less centralized, primary care facilities.

(ii) Temporal Artifacts: The inclusion of data spanning from 1999-2023 introduces historical bias. Earlier records (pre-2008) often lack structured diagnostic fields, potentially affecting the accuracy of longitudinal analysis.

(iii) Data Specificity Constraints: The privacy risk estimates primarily derive from a renal disease database. While China has established EMR documentation guidelines, structural variations between the CRDS and other data networks may require adjustments in the protocol to ensure applicability to broader datasets. The methodology should be refined to account for such differences to be applicable to a wider range of diseases and settings.

(iv) Over-masking Artifacts: The reliance on regular expressions for data desensitization may lead to occasional over-masking, where non-sensitive data is incorrectly masked. This can hinder the utility of certain records. Future iterations of the system will explore the integration of advanced natural language processing (NLP) models, such as transformer-based architectures, to improve the balance privacy protection and data retention.

### Clinical Implications

The study identifies two critical exposure pathways that require further attention for improving the privacy protection of psychiatric data:

(i) Covert Leakage: Some sensitive psychiatric data may indirectly surface through laboratory results or clinical parameters. For instance, specific medication metabolites (e.g., olanzapine metabolite levels) or therapeutic procedures (e.g. repetitive transcranial magnetic stimulation parameters) may contain identifiable information. Such indirect pathways highlight the need for more comprehensive masking strategies that protect all forms of sensitive data.

(ii) Associative Inference: Partially redacted identifiers, such as “PSY***” hospitalization codes, could still be indirectly linked to psychiatric hospital databases, potentially revealing sensitive patient information. To address this risk,we propose a tri-level de-identification strategy: Level 1: Direct sensitive information masking(e.g., psychiatric disorders, medications, diagnosis codes);Level 2: Contextual inference prevention by eliminating any identifiable metadata associated with sensitive terms; Level 3: Cross-database linkage disruption, ensuring that even partially redacted records cannot be re-identified when combined with other datasets.

This multi-layered de-identification approach ensures that not only direct identifiers but also context-dependent inferences are adequately addressed, thus providing a more robust mechanism for safeguarding sensitive psychiatric data. Furthermore, these strategies align with compliance requirements under China’s Personal Information Protection Law (PIPL), ensuring both privacy protection and data utility in healthcare settings.

### Future Directions

Building on the OHDSI Common Data Model, the following steps are prioritized: (i)Developing multilingual privacy pipelines that ensure precise mapping between Chinese and English terms; (ii)Creating dynamic risk assessment models quantifying privacy exposure coefficients across scenarios, such as clinical research, insurance underwriting, employment screening; (iii)Implementing federated learning frameworks to enable cross-institutional analysis while maintaining data localization.

## Conclusion

The EPPDI protocol provides an innovative solution to the challenges of privacy protection in Chinese electronic medical records (EMRs), particularly in the context of sensitive mental health information. By resolving terminological ambiguity and adapting to unstructured narratives (e.g., implicit diagnoses in clinical course descriptions), this system achieves high precision and recall rates in identifying psychiatric disorder-related data while preserving data utility.

This framework not only addresses the specific needs of China’s Personal Information Protection Law (PIPL) but also provides a broader technical paradigm for data privacy protection within the Chinese healthcare system, particularly for sensitive mental health data. By combining advanced natural language processing (NLP) with traditional privacy-preserving strategies, EPPDI ensures compliance with stringent legal standards while maintaining the usability of critical medical data for clinical and research purposes.

Looking ahead, the future of privacy protection in healthcare will depend on the continued development of adaptive de-identification tiers that balance privacy preservation with the evolving needs of medical research. Future work should focus on refining these methods by integrating machine learning techniques to enhance the system’s ability to dynamically adjust masking depth based on the specific objectives of each study. Additionally, expanding the system’s applicability to other medical domains, such as oncology or infectious diseases, will further demonstrate the scalability and versatility of the EPPDI protocol in diverse healthcare settings. Furthermore, leveraging emerging technologies like federated learning could facilitate cross-institutional data sharing without compromising privacy, thus enabling broader collaboration in medical research while ensuring patient confidentiality.

## Data Availability

The datasets supporting the findings of this study are available from the corresponding author upon reasonable request. The hospital data cannot be made publicly available due to its sensitive nature.

## Declarations

### Author contributions

Mengchun Gong, Endi Cai and Minghao Que wrote the main manuscript text and Chaoliu, Yiling Zhao, Zihao Ouyang provided some technical support to facilitate our data extraction work and also verified and reviewed our data. Wenzhao Shi, Mengchun Gong and Sheng Nie guided the direction of the article. All authors reviewed the manuscript.

## Abbreviations

EMR: Electronic medical records
PD: Psychiatric Disorders
RWE: Real-world evidence
CRDS: Chinese renal disease data system
EPPDI: Extraction Protocol of Psychiatric Disorders Information
NLP: Natural language processing
TP: True positives
FP: False positives

## References

1. Huang, Y., et al., Prevalence of mental disorders in China: a cross-sectional epidemiological study. Lancet Psychiatry, 2019. 6(3): p. 211–224.

2. World Health, O., International statistical classification of diseases and related health problems. 10th revision, Fifth edition, 2016 ed. 2015, Geneva: World Health Organization.

3. Altawy, R. and A.M. Youssef, Security Tradeoffs in Cyber Physical Systems: A Case Study Survey on Implantable Medical Devices. IEEE Access, 2016. 4: p. 959–979.

4. Hadzic, M., M. Chen, and T.S. Dillon. Towards the Mental Health Ontology. in 2008 IEEE International Conference on Bioinformatics and Biomedicine. 2008.

5. Basil, N.N., et al., Health Records Database and Inherent Security Concerns: A Review of the Literature. Cureus, 2022. 14(10): p. e30168.

6. Chmielewski, M.R., ONC Releases Updated Guide to Privacy and Security of Electronic Health Information.

7. Papoutsi, C., et al., Patient and public views about the security and privacy of Electronic Health Records (EHRs) in the UK: results from a mixed methods study. BMC Medical Informatics and Decision Making, 2015. 15(1): p. 86.

8. Gariepy-Saper, K. and N. Decarie, Privacy of electronic health records: a review of the literature. J Can Health Libr Assoc, 2021. 42(1): p. 74–84.

9. Bondre, A., S. Pathare, and J.A. Naslund, Protecting Mental Health Data Privacy in India: The Case of Data Linkage With Aadhaar. Glob Health Sci Pract, 2021. 9(3): p. 467–480.

10. Personal Information Protection Law of China. 2021. 000(004): p. P.28-36.

11. Fitzgerald, J. The 10 Biggest Data Breaches Of 2022. Available from: https://healthitsecurity.com/news/the-10-biggest-healthcare-data-breaches-of-2020-so-far.

12. China, N.H.C.o.t.P.s.R.o., Technical specification of hospital information platform based on electronic medical record. 2014.

13. Wang, Z., Data integration of electronic medical record under administrative decentralization of medical insurance and healthcare in China: a case study. Isr J Health Policy Res, 2019. 8(1): p. 24.

14. Rajput, A.R., et al., EACMS: Emergency Access Control Management System for Personal Health Record Based on Blockchain. 2019.

15. Shahnaz, A., U. Qamar, and A.J.I.A. Khalid, Using Blockchain for Electronic Health Records. 2019. PP(99): p. 1–1.

16. Xu, J., et al., Healthchain: A Blockchain-Based Privacy Preserving Scheme for Large-Scale Health Data. IEEE Internet of Things Journal, 2019. 6(5): p. 8770–8781.

17. Ren, H., et al., Privacy-Enhanced and Multifunctional Health Data Aggregation under Differential Privacy Guarantees. Sensors (Basel), 2016. 16(9).

